# Development and validation of a dynamic risk stratification tool for predicting multidrug-resistant bacterial infections in ICU patients: A clinical prediction model and web-based calculator

**DOI:** 10.64898/2026.05.23.26353927

**Authors:** Lingling Ye, Binghang Lyu, Qijuan Yang, XiuHuan Mou, Rajitra Nawawonganun, Wongsa Laohasiriwong

**Author notes:** These authors contributed equally to this work.

## Abstract

**Background:** Multi-drug resistant Bacterial (MDRB) Infections in the intensive care units (ICUs) substantially elevate patient mortality, prolong hospital stays, and impose heavy healthcare cost burdens. Existing predictive models for ICU-acquired MDRB infection predominantly focus on static admission-risk assessment, lacking the capacity to leverage longitudinal treatment data for dynamic risk re-stratification during the ICU stay. Meanwhile, most models suffer from poor clinical interpretability, overreliance on hard-to-collect biomarkers, or absence of deployable clinical tools, limiting real-world translation. Therefore, there is an urgent need to develop a parsimonious, interpretable tool based on routine cumulative data to guide timely intervention. This study aimed to develop a interpretable model with a web calculator to improve clinical applicability.

**Methods:** In this study, we conducted a retrospective analysis of ICU inpatients at the First Affiliated Hospital of Dali University between January 1, 2023, and January 1, 2026. Using the create Data Partition function in R software (random seed = 42), the dataset was stratified and divided into a training group and a validation group in a 7:3 ratio. Feature selection was performed using the Boruta algorithm to validate variable rationality. A multivariable logistic regression model was constructed and visualized as a nomogram, and its performance was compared with six machine learning algorithms (Random Forest, XG Boost, Neural Network, etc.). Model validation was conducted using receiver operating characteristic curves (ROC), Decision Curve Analysis (DCA), and SHAP value interpretation. Finally, an online R Shiny calculator was developed based on the final model.

**Results:** A total of 3,631 patients were enrolled and divided into a training group (n=2,543) and a validation group (n=1,088) using stratified random sampling. Five independent predictors were identified in the training group, which were hypertension combined with diabetes, antibiotic types, ventilator days, urinary catheter days, and PCT abnormality times. The Logistic regression model achieved an AUC of 0.772 (95%CI: 0.733-0.812) in the validation group, outperforming XG Boost (0.763) and Random Forest (0.703). The model demonstrated excellent calibration (Hosmer-Leme show χ² = 1.94, P = 0.9829) and positive net clinical benefit across threshold probabilities of 0%–40%. SHAP analysis aligned with regression-derived variable importance rankings, confirming predictor contributions. An open-access online calculator was successfully deployed (https://dongfangshao666.shinyapps.io/MDR_shiny2/), enabling real-time individualized risk stratification at the bedside.

**Conclusion:** This study developed and validated a dynamic, interpretable multi-drug-resistant bacterial infection risk prediction model requiring only five routinely collected clinical indicators. The model balances robust predictive performance with high transparency, overcoming key limitations of prior tools. The accompanying web calculator supports dynamic risk reassessment throughout the ICU stay, facilitating precise antimicrobial stewardship, targeted infection control interventions, and optimized resource allocation, bridging the gap between statistical modeling and frontline clinical decision-making.

## Introduction

Multidrug-resistant bacterial (MDRB) infection has become a major global public health challenge, especially in the intensive care unit (ICU) setting, where they are a major concern due to the increased mortality rate, prolonged hospital stay, sharp increase in medical costs and excessive consumption of medical resources it causes [1,2]. Some studies found that patients with hospital-acquired MDRB infections in the ICU had an average 13% longer ICU stay and an average 26% higher total hospitalization cost compared to patients with infections caused by non-multidrug-resistant bacteria. [3]. Therefore, early and accurate identification of high-risk patients with MDR infection is crucial for guiding the rational use of empirical antibacterial drugs, timely implementation of contact isolation, and optimization of infection control strategies [4,5].

Although several predictive models for multidrug-resistant infections in ICU patients have been reported in the past, existing studies often have limitations. Some models rely excessively on complex machine learning algorithms such as random forests and deep learning; while their predictive performance is acceptable, they lack clinical interpretability and struggle to reveal the underlying associations between variables and outcomes, hindering clinicians’ trust and adoption [6]. Other models include an excessive number of variables or rely on biochemical indicators that are difficult to obtain, making them hard to apply quickly in the fast-paced clinical setting [7,8]. Furthermore, the vast majority of models remain at the stage of statistical charts, lacking accompanying visual interactive tools or online calculators, which greatly limits the translation and application of predictive models in clinical practice [9,10].

This study aims to construct a variable-reduced, highly interpretable MDR infection risk prediction model based on real-world data from the First Affiliated Hospital of Dali University. We achieve this by performing objective feature selection using the Boruta algorithm, combining it with classical logistic regression, and comparing its performance against Random Forest (RF) and XG Boost algorithms. More importantly, we will further transform this model into an interactive online calculator based on the R Shiny framework, with the goal of bridging the gap between statistical modeling and bedside clinical decision support.

## Material and methods

### Study design and ethical approval

This retrospective study was conducted in the 54–bed ICU of a 2500–bed tertiary general hospital in Dali, China, covering the period from January 2023 to January 2026. A total of 3631 patients were screened, and the final analysis focused on those who developed multidrug–resistant bacterial infections during ICU stay. To ensure the robustness of the model development, we employed the create Data Partition function in R with a fixed random seed (seed=42) to perform stratified random sampling, dividing the dataset into a training group (70%, n=2543) and a validation group (30%, n=1088) while maintaining a consistent MDR positivity rate in both cohorts. Internal validation was conducted using 5-fold cross-validation and Bootstrap resampling (1000 iterations) to assess model stability and optimism. Moreover, to evaluate the performance of the logistic regression model, we conducted a comparative analysis against six state-of-the-art machine learning classifiers (Neural Network, XG Boost, Naive Bayes, Decision Tree, Random Forest, and Support Vector Machine), all trained and validated under identical data partitioning conditions.

This study was approved by the Ethics Committee of the First Affiliated Hospital of Dali University (Approval No. DFY20260429003) and the Khon Kaen University Ethics Committee for Human Research (Approval No. HE692040).

### Study population and outcome definition

Multidrug-resistant infections were defined as non-susceptibility to at least one agent in three or more antimicrobial categories [11]. The study population comprised of all adult patients admitted to the ICU during the study period. Patients were excluded if they met any of the following criteria: (i) length of hospital stay <48 hours; (ii) known immunodeficiency; or (iii) confirmed bacterial colonization at admission. To prevent data duplication, only the first isolation of a specific bacterial strain from a single specimen type per patient was included in the analysis.

The primary outcome was the occurrence of an MDR infection during the ICU stay, confirmed by the isolation of pathogens resistant to three or more antimicrobial classes from clinical specimens. To enhance clinical interpretability and facilitate bedside application, continuous variables were transformed into categorical variables. For instance, the number of antibiotic types administered was categorized as ≤1, 2, 3, or >3; durations of mechanical ventilation and urinary catheterization were dichotomized or trichotomized using a 14-day cutoff.

### Data collection and candidate variables

Clinical data were extracted from the electronic medical record system, covering 19 candidate variables across four domains: demographics, comorbidities, hospitalization indicators, and infection–related interventions. Laboratory and medication data were recorded according to standard clinical protocols.

### Statistical analysis and model development

Patients with missing data in key variables were excluded from the analysis. Continuous variables were assessed for normality using the Shapiro–Wilk test; non-normally distributed variables were presented as median (interquartile range, IQR) and compared using the Mann–Whitney Utest, while categorical variables were presented as frequencies (%) and compared using the chi–square (χ²) test.

Feature selection was performed using the Boruta algorithm (R package Boruta) with 100 random forest runs and a significance threshold of P< 0.01, comparing original attributes against shadow attributes generated via random permutation. To validate the linearity assumption for continuous variables, Restricted Cubic Spline (RCS) analysis with three knots was applied. Notably, the non-linearity test for PCT abnormality times yielded a P-value of 0.146, supporting its inclusion as a linear continuous variable in the final model.

A multivariable logistic regression model was constructed based on the selected predictors, and model coefficients were used to develop a visualized nomogram. To benchmark its performance, six alternative machine learning algorithms were implemented, which were Random Forest (RF), Extreme Gradient Boosting (XG Boost), Neural Network, Naïve Bayes, Decision Tree, and Support Vector Machine (SVM). Hyperparameter tuning was conducted for each model using grid search combined with 5–fold cross–validation. Specifically, we performed grid search for XG Boost and SVM (testing linear vs. radial basis function kernels), optimized *m-try* and *n-tree* for random forest, tuned the complexity parameter (*cp*) for decision trees, and adjusted size and decay for neural networks. Default parameters were retained for naïve Bayes with discretization strategies.

Model performance was rigorously evaluated in the internal validation group. Discrimination was assessed using the area under the receiver operating characteristic curve (AUC) with 95% confidence intervals calculated via the DeLong method. Calibration was evaluated using the Hosmer–Lemeshow goodness–of–fit test and bootstrap–corrected calibration plots (1,000 resamples). Clinical utility was quantified using Decision Curve Analysis (DCA). Additionally, SHapley Additive exPlanations (SHAP) values were computed based on the XG Boost model to cross–validate predictor importance rankings and linear contributions from a machine learning perspective. All statistical analyses were performed using R software (version 4.3.0), and the final predictive model was deployed as a web–based calculator using the R Shiny framework.

### Ethics statement

Due to the retrospective nature of the study and the use of de-identified data, the requirement for informed consent was waived by both ethics’ committees. All methods were carried out in accordance with relevant guidelines and regulations.

## Results

### Patient Characteristics and Baseline Data

A total of 3,631 ICU patients were included in this study, comprising 2,328 males (64.11%) and 1,303 females (35.89%). The median age was 59 years, with 48.97% of patients aged ≥60 years. Regarding comorbidities, the prevalence of hypertension was 35.03%, type 2 diabetes mellitus was 15.29%, and the coexistence of both conditions was 9.91%. The most frequent admission diagnoses were respiratory system diseases (68.71%) and brain disorders (33.65%).

For treatment-related variables, the median length of stay (LOS) in the ICU was 15 days (interquartile range [IQR]: 3–305), and 43.10% of patients underwent surgical intervention. Regarding antimicrobial exposure, 53.59% of patients received antibiotics for 0–14 days, with 26.96% receiving more than three antibiotics concurrently. Invasive device utilization was common, with median ventilator days of 4 (IQR: 0–298), median urinary catheter days of 9 (IQR: 0–302), and median central venous catheter days of 7 (IQR: 0–209). The median frequency of abnormal procalcitonin tests was 1 (IQR: 0–28). Group comparisons between the training and validation sets were performed using the Mann–Whitney Utest for continuous variables and the χ² test for categorical variables. All variables were well-balanced between the two groups (P> 0.05), except for the combination application of antibiotics (P= 0.044) (Table 1).

**Table 1.**
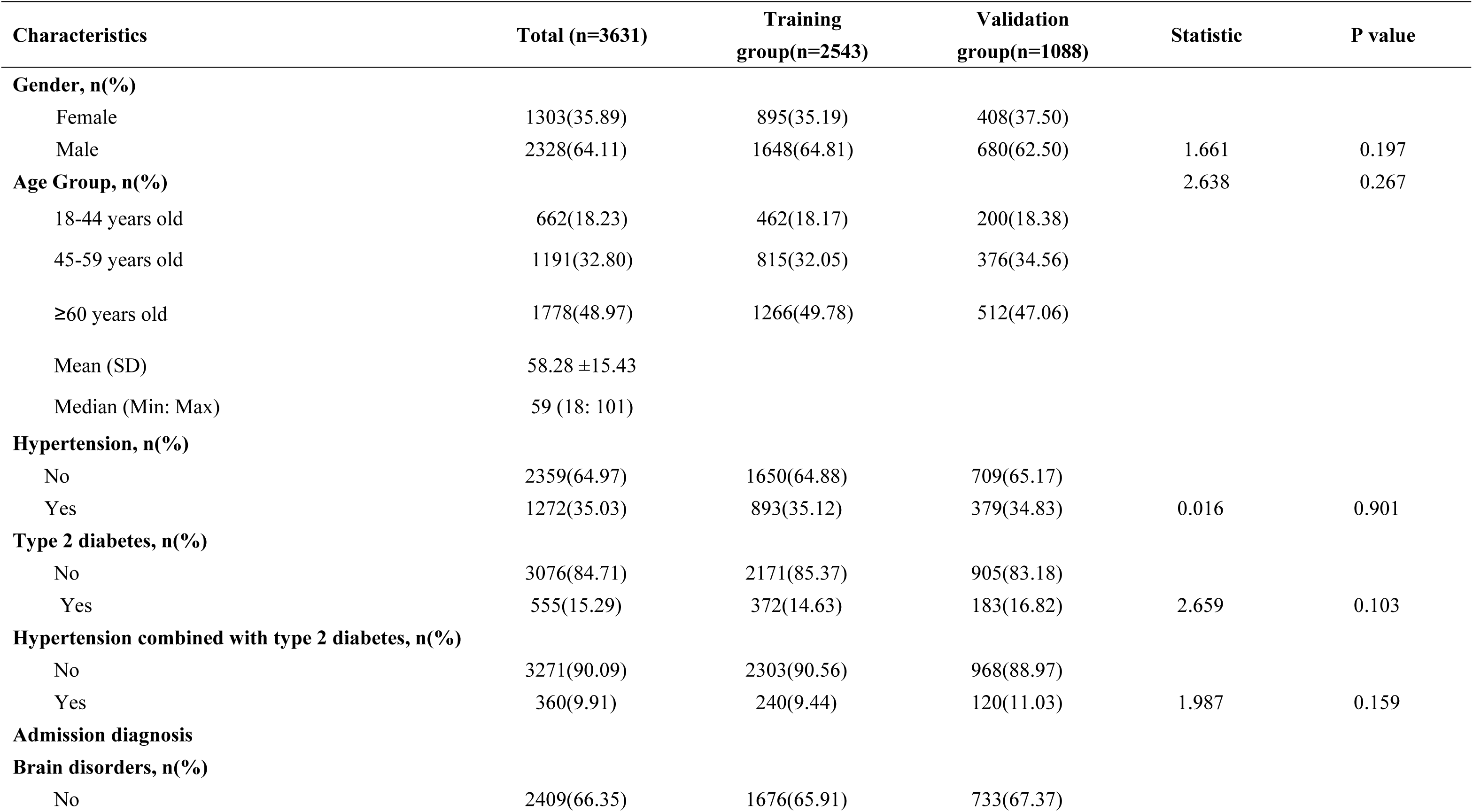
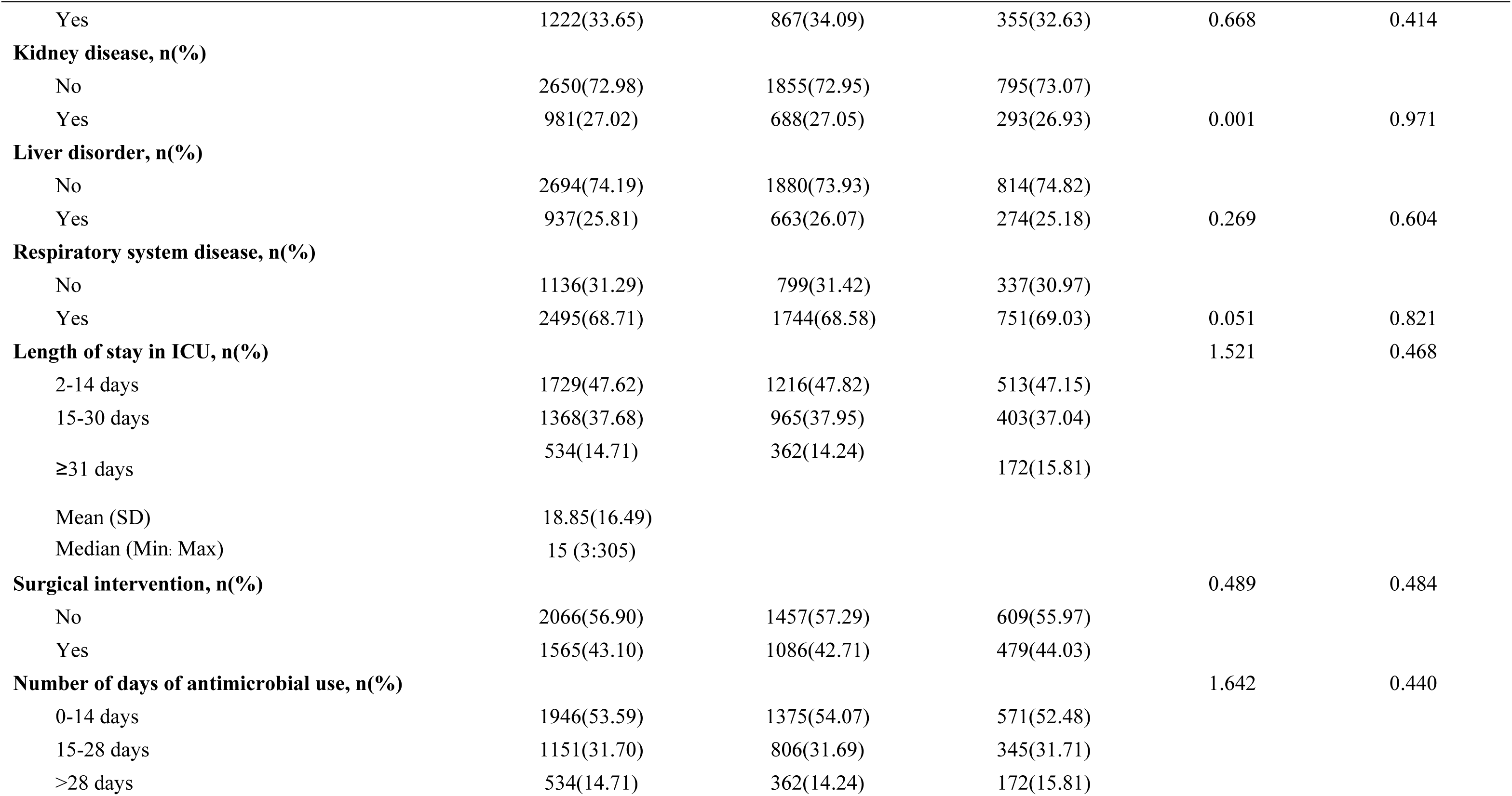
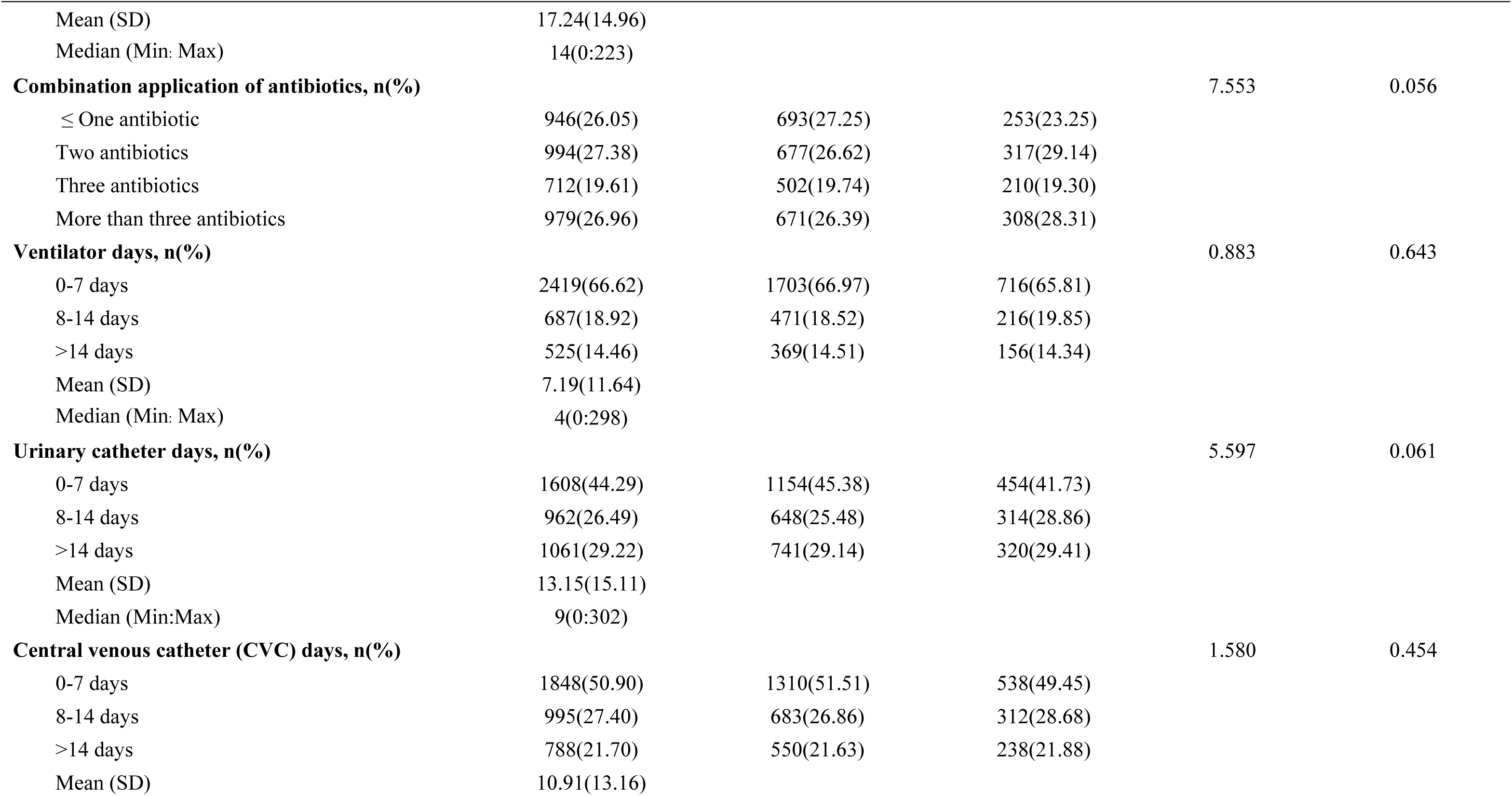
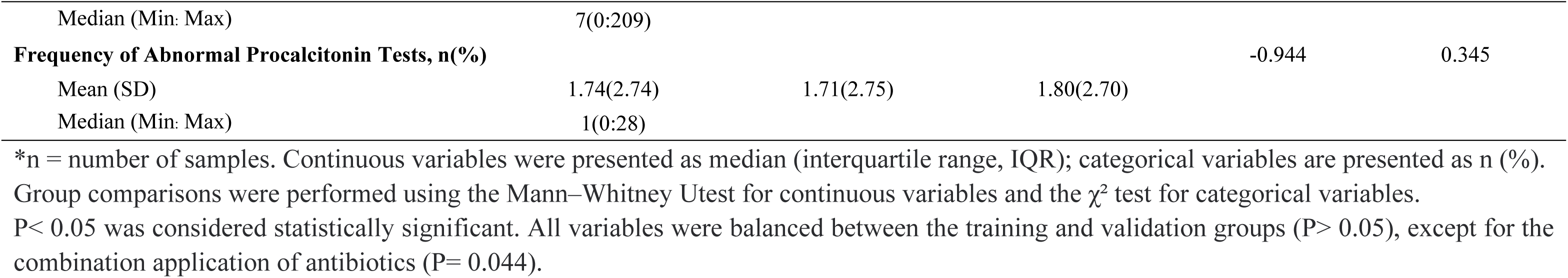
Baseline characteristics of ICU patients in the training and validation groups.

### Variable screening and model construction

We used the Boruta algorithm to conduct a full feature screening of all candidate variables in the original data to determine which variables had statistically significant explanatory power for the outcome. Based on the ranking, the top predictive factors (highlighted in green) primarily consist of treatment-related exposures and invasive device utilization days. Specifically, these include: Ventilator Days (Duration of mechanical ventilation), Antibiotic Days (Duration of antibiotic therapy), CVC days (Duration of central venous catheterization), Urinary catheter Days (Duration of urinary catheterization), LOS (Length of stay in ICU), Combined Antibiotics(Combination of antibiotic types used), and m-PCR Count(Frequency of abnormal procalcitonin tests) (Fig 1).

**Fig 1.**
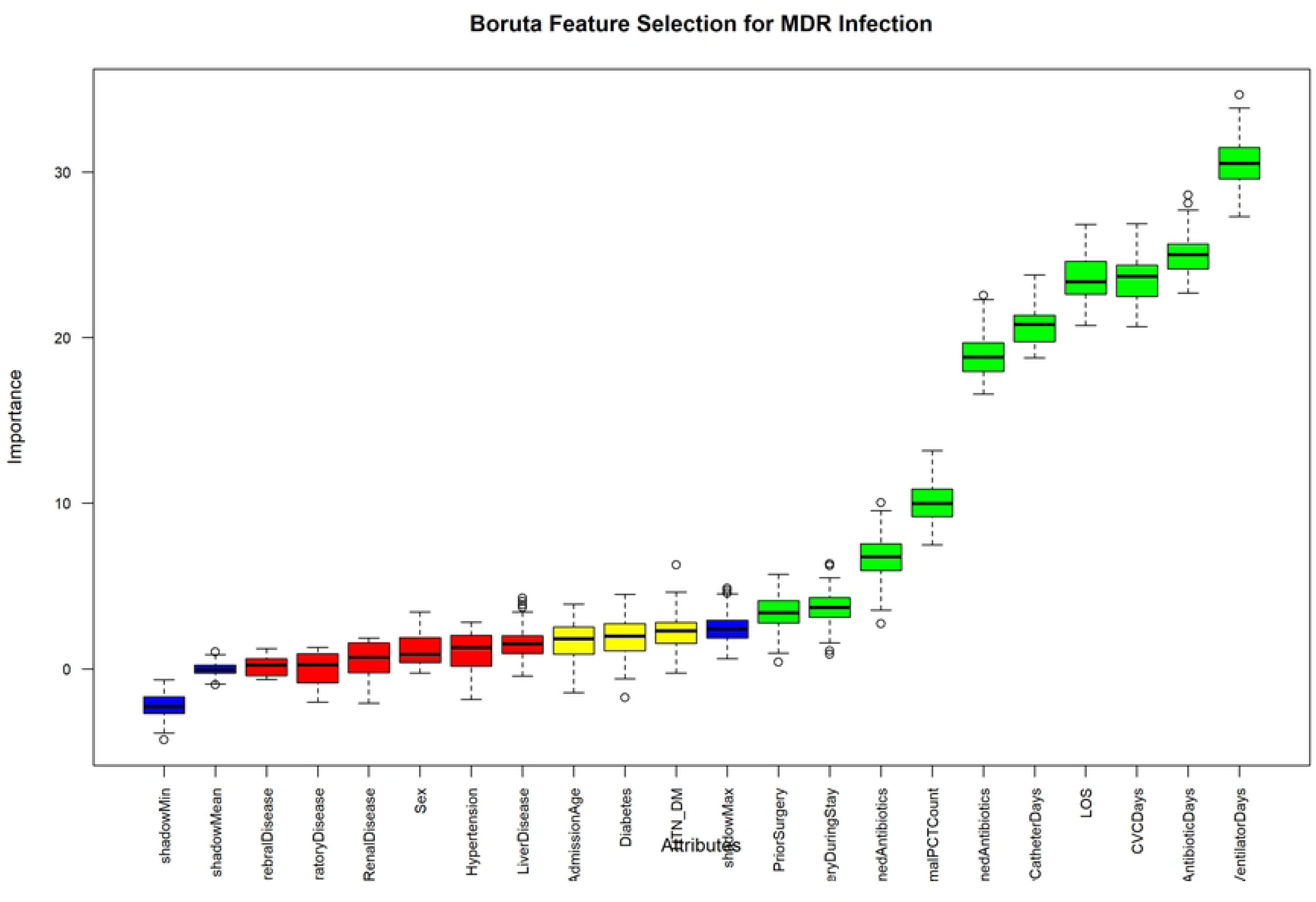
Feature selection using the Boruta algorithm. Variables with importance scores significantly higher than randomized shadow attributes (green) were retained, while those without significance(red) were excluded.

Subsequently, univariate and multivariate logistic regression analyses were performed on the features identified by the Boruta algorithm. Considering the trade-off between model parsimony and predictive performance, five independent predictors were ultimately selected and incorporated into the final prediction model: hypertension combined with type 2 diabetes, combination antibiotic therapy, ventilator days, urinary catheter days, and frequency of abnormal procalcitonin tests (Table 2). Based on these predictors, a visualization nomogram was constructed using the training group to facilitate individualized risk prediction (Fig 2).

**Table 2.**
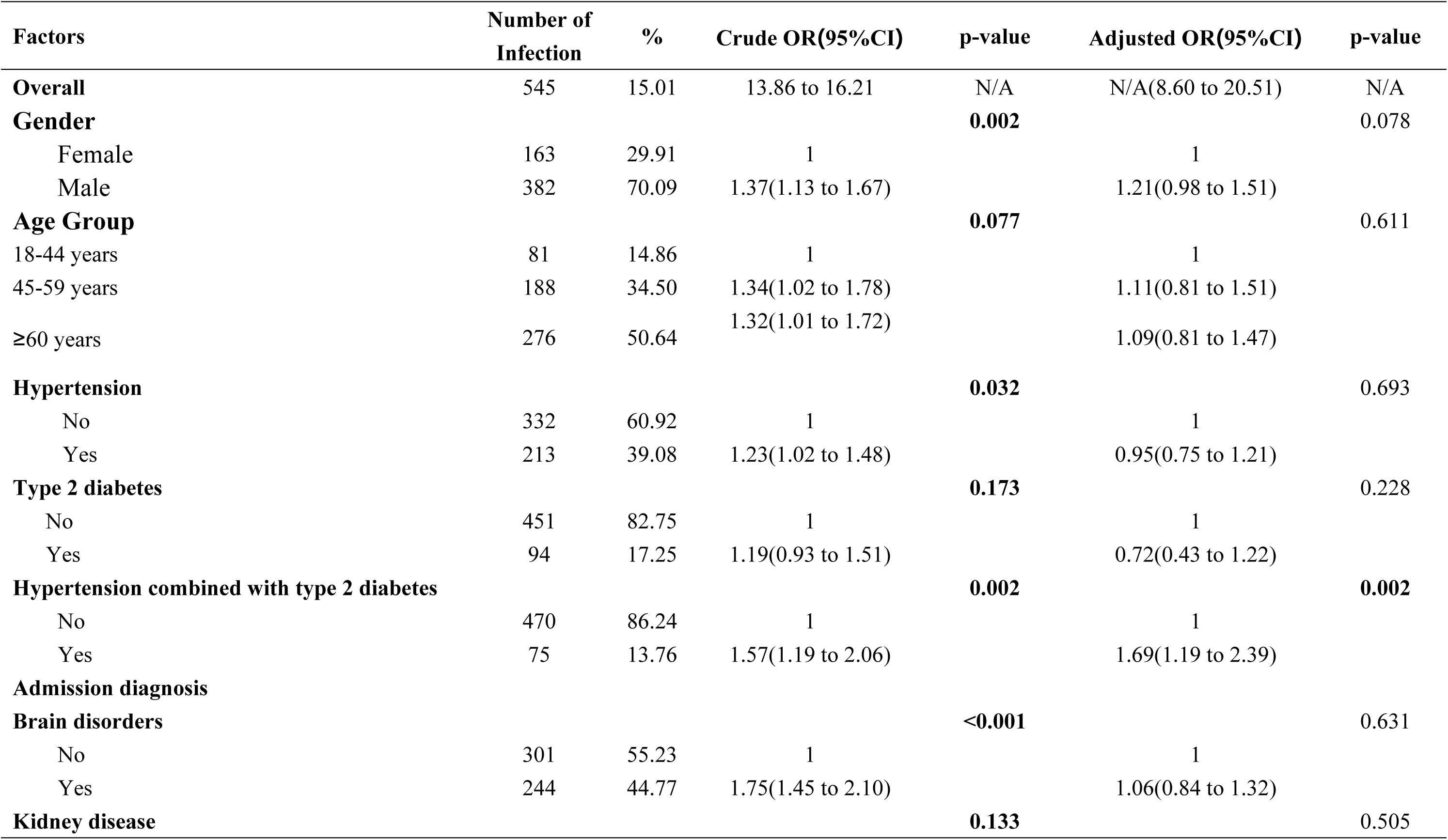
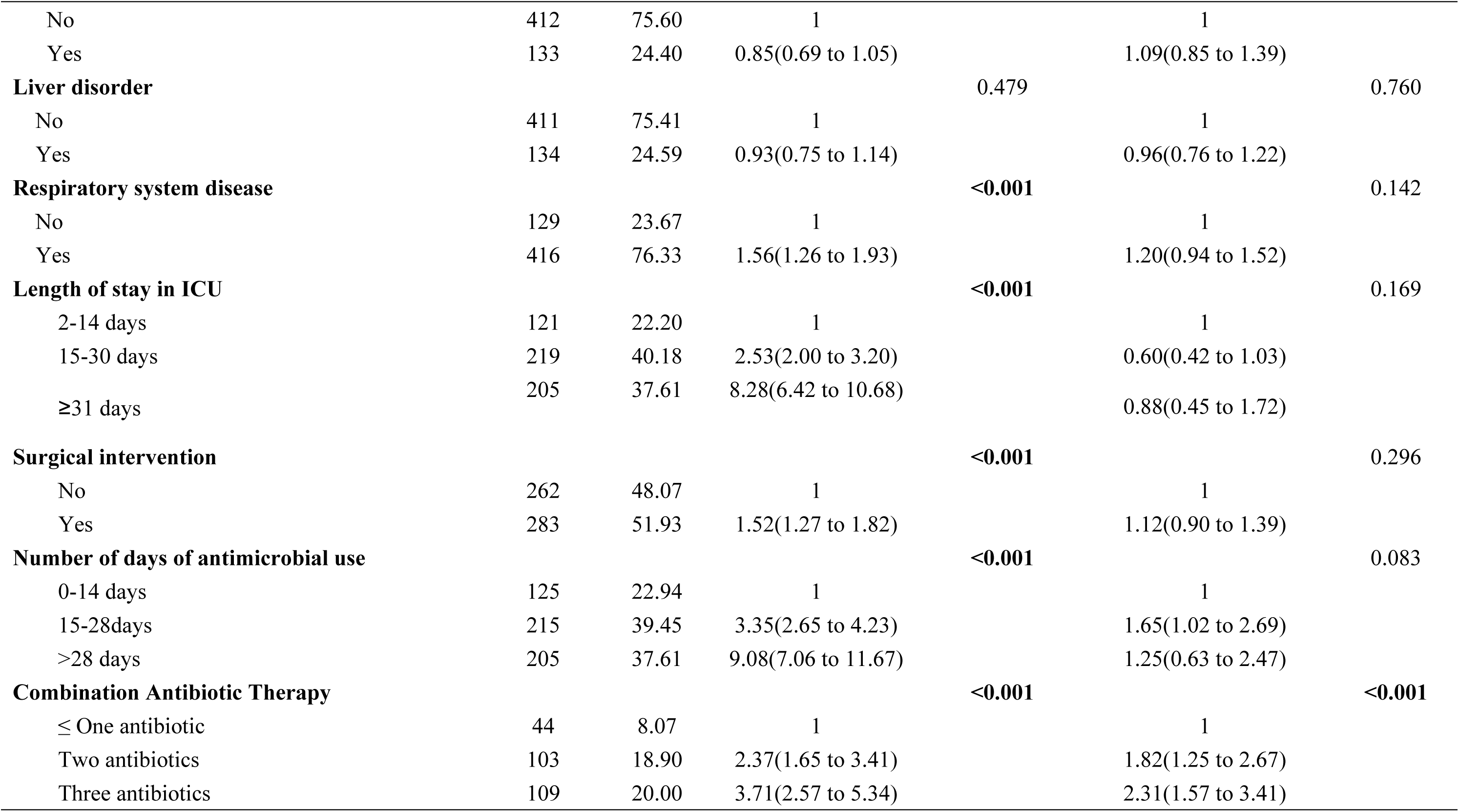
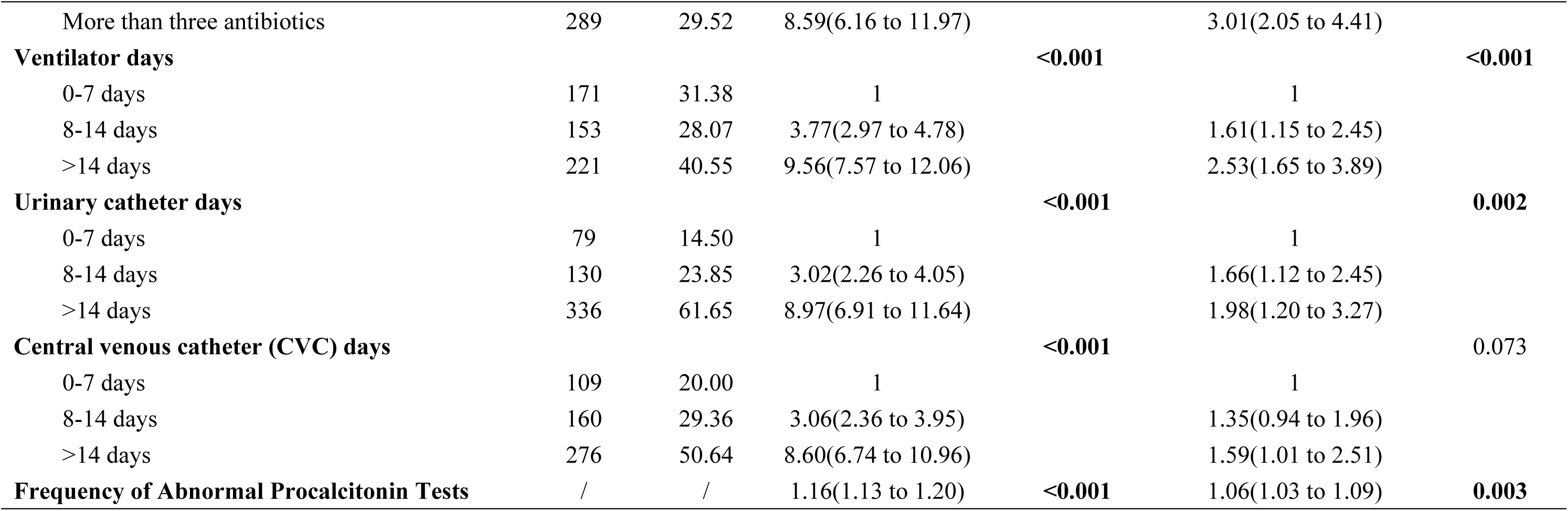
Logistic regression analysis of predictive factors for MDRB infection in ICU patients.

**Fig 2.**
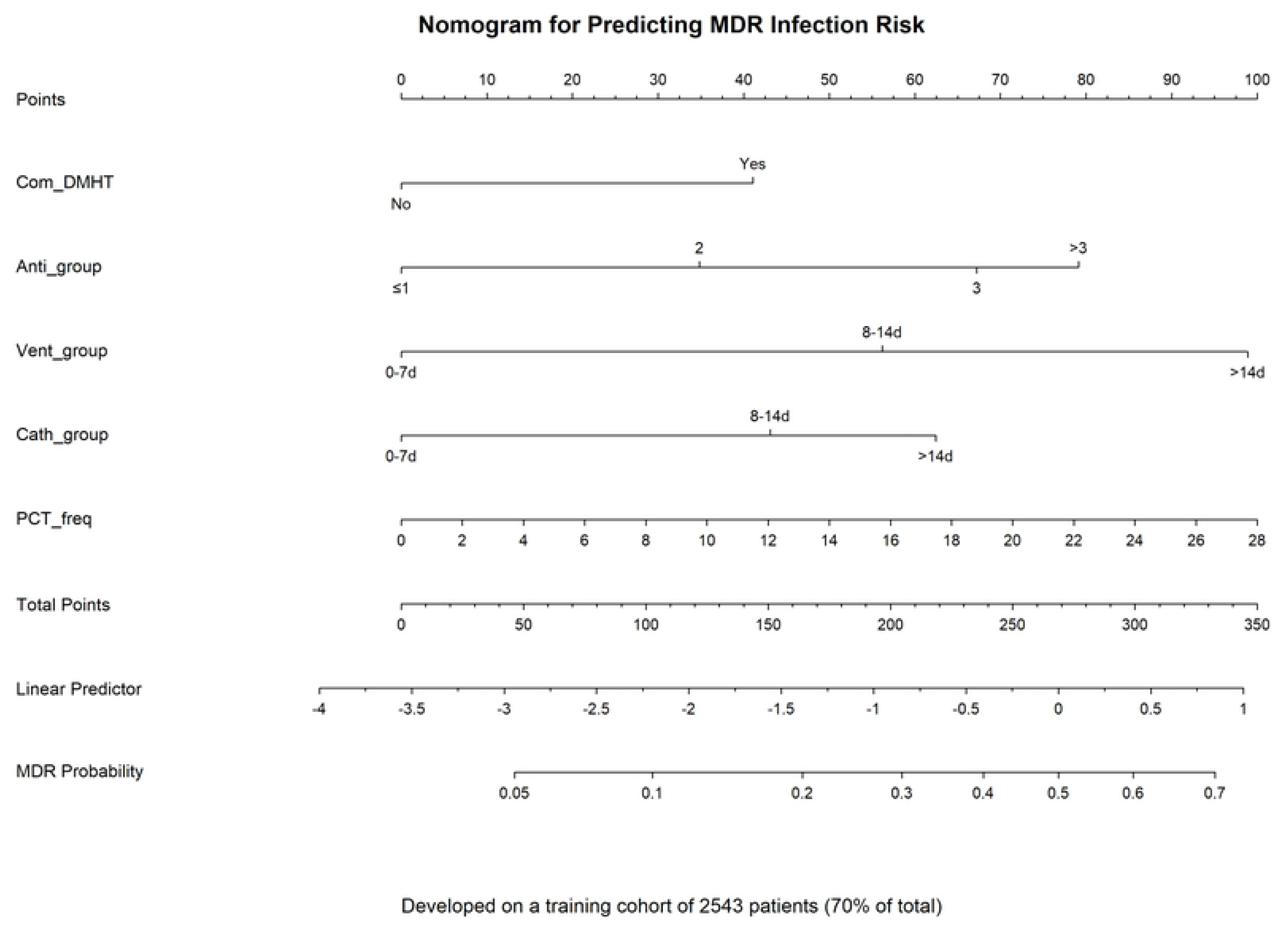
Nomogram for predicting MDR infection risk developed using the training group. The nomogram converts the multivariable logistic regression model into a visual scoring system. Each predictor corresponds to a point scale (top axis). Total points are summed and mapped to the bottom axis to estimate the individual probability of developing an MDR infection during the ICU stay.

### Model Validation and Performance Evaluation

In the independent validation group (n=1088), the logistic regression model demonstrated robust discriminatory ability with an AUC of 0.772 (95% CI: 0.733–0.812), outperforming XG Boost (AUC=0.763, 95%CI: 0.723-0.803) and Random Forest (AUC=0.703, 95%CI: 0.658-0.748) (Fig 3). Using the Youden index, the optimal cutoff value was determined to be 0.2176, yielding a sensitivity of 60.7% and specificity of 80.2%. Patients were stratified into low-risk (<10%), medium-risk (10%–30%), and high-risk (>30%) groups, with actual MDR infection rates of 5.99%, 15.29%, and 42.78%, respectively. Notably, the model achieved a high Negative Predictive Value (NPV) of 92.1%. The performance comparison of the tuned logistic regression model against six alternative machine learning algorithms is summarized in Table 3.

**Table 3.** Performance evaluation metrics of various tuned models for predicting MDR infection.

| Models (Tuned) | AUC | AUC 95% CI | *P* value | Cutoff Value | Precision (%) | Sensitivity (%) | Specificity (%) | Accuracy (%) | F1 Score | G-mean | Brier Score |
| --- | --- | --- | --- | --- | --- | --- | --- | --- | --- | --- | --- |
| **Logistic Regression** | 0.772 | 0.732–0.811 | <0.001 | 0.218 | 35.1 | 60.7 | 80.2 | 77.3 | 0.445 | 0.698 | 0.108 |
| **Neural Network** | 0.768 | 0.728–0.808 | <0.001 | 0.168 | 35.0 | 63.8 | 79.1 | 76.8 | 0.452 | 0.711 | 0.109 |
| **XGBoost** | 0.767 | 0.726–0.807 | <0.001 | 0.233 | 37.2 | 57.7 | 82.8 | 79.0 | 0.452 | 0.691 | 0.110 |
| **Naive Bayes** | 0.740 | 0.694–0.787 | <0.001 | 0.097 | 30.8 | 68.7 | 72.8 | 72.2 | 0.425 | 0.707 | 0.164 |
| **Decision Tree** | 0.724 | 0.682–0.767 | <0.001 | 0.085 | 29.6 | 67.5 | 71.7 | 71.0 | 0.411 | 0.695 | 0.114 |
| **Random Forest** | 0.709 | 0.666–0.753 | <0.001 | 0.001 | 27.2 | 66.9 | 68.4 | 68.2 | 0.387 | 0.676 | 0.128 |
| **Support Vector Machine** | 0.669 | 0.621–0.717 | <0.001 | 0.138 | 35.8 | 42.3 | 86.6 | 80.0 | 0.388 | 0.605 | 0.117 |

**Fig 3.**
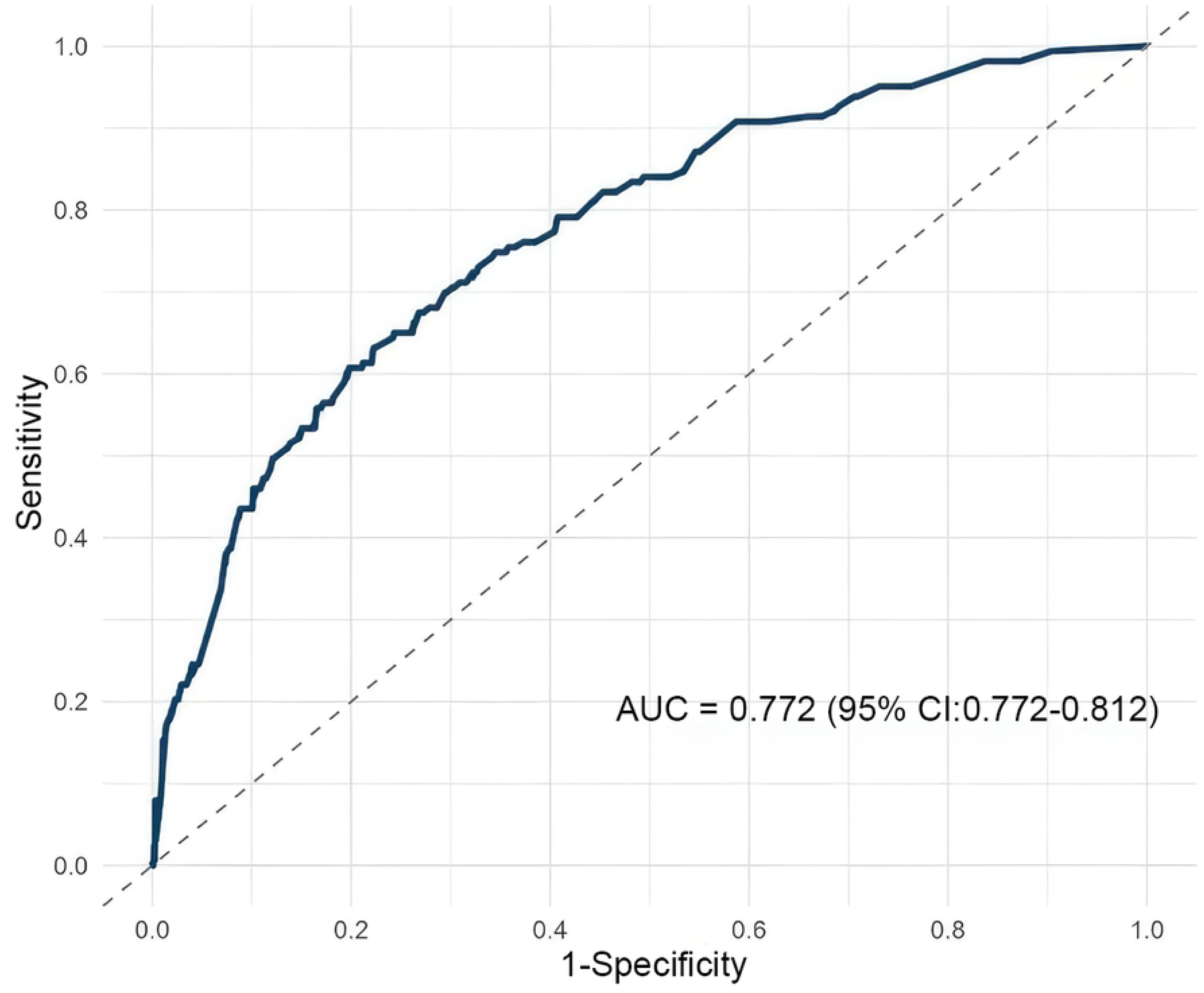
ROC curve of the prediction model for the validation Group. Receiver Operating Characteristic (ROC) curve. The area under the curve (AUC) was 0.772 (95% CI: 0.733–0.812), indicating moderate discriminative ability. The dashed line represents the performance of a random classifier.

To visually compare the discriminative abilities, the Receiver Operating Characteristic (ROC) curves of all seven models are presented in Fig 4. As shown in this figure, the tuned Logistic Regression model demonstrated comparable discriminatory power to the top-performing XG Boost and Neural Network models, with all three models achieving an AUC above 0.76. In contrast, the Support Vector Machine exhibited the poorest performance.

**Fig 4.**
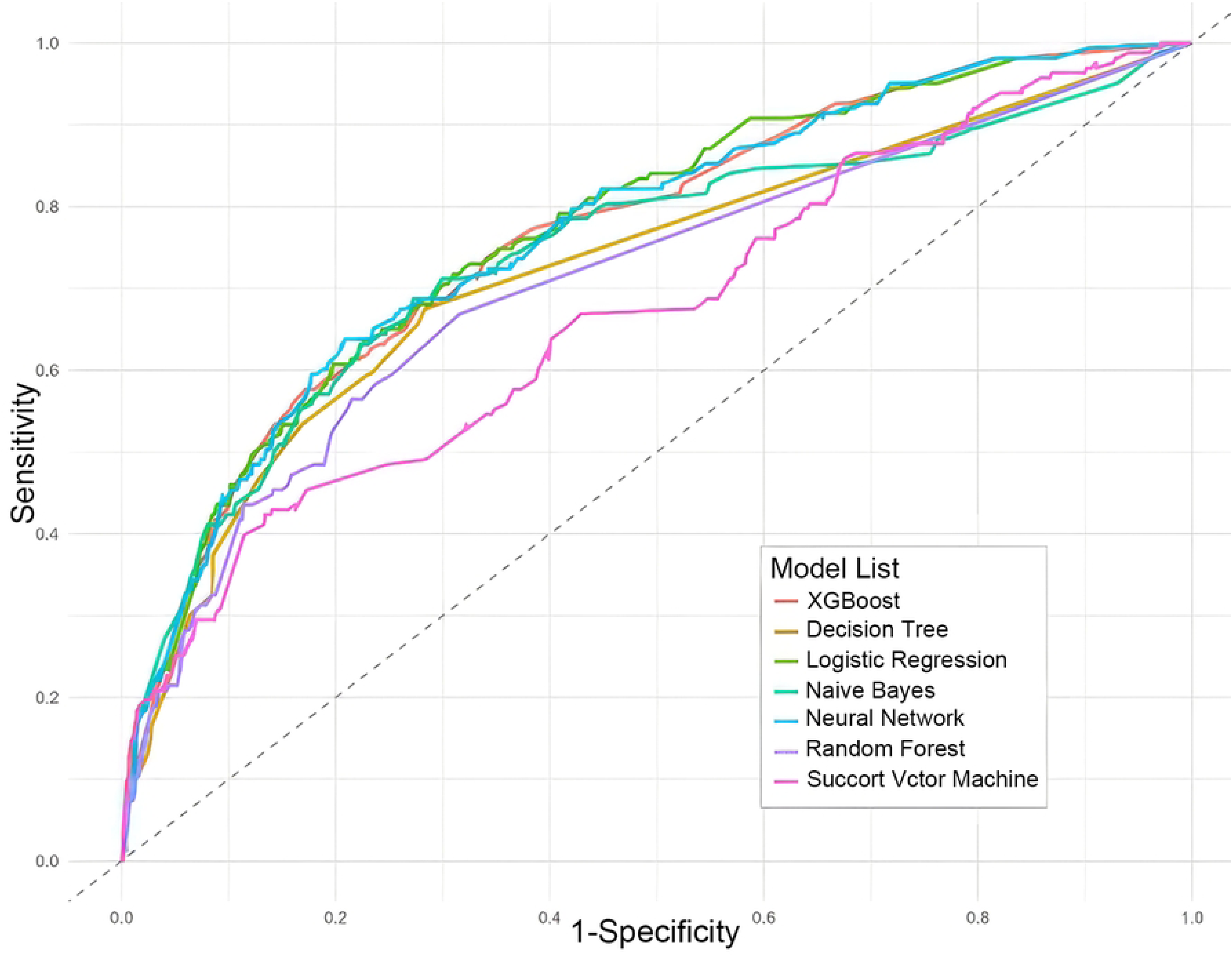
ROC curves of the seven tuned machine learning models for predicting MDR infection. The ROC curves compare the performance of XG Boost (red), Decision Tree (yellow), Logistic Regression (light green), Naive Bayes (dark green), Neural Network (cyan), Random Forest (purple), and Support Vector Machine (pink) in predicting multidrug-resistant (MDR) infections. The area under the curve (AUC) reflects the discriminatory ability of each model, with curves closer to the top-left corner indicating better performance. The diagonal dashed line represents a random classifier with an AUC of 0.5.

The model calibration was excellent, as evidenced by the Hosmer-Lemeshow test (X^2^=1.94, P=0.9829) and the close alignment of the calibration curve with the ideal line (Fig 5). Five-fold cross-validation yielded an average AUC of 0.78±0.03, consistent with the independent validation results. Bootstrap resampling (1000 iterations) revealed a minimal optimism of 0.005, indicating negligible overfitting. Sensitivity analysis using different random seeds (123, 456, 999) showed minimal AUC fluctuation (0.763–0.787), confirming the model’s stability.

**Fig 5.**
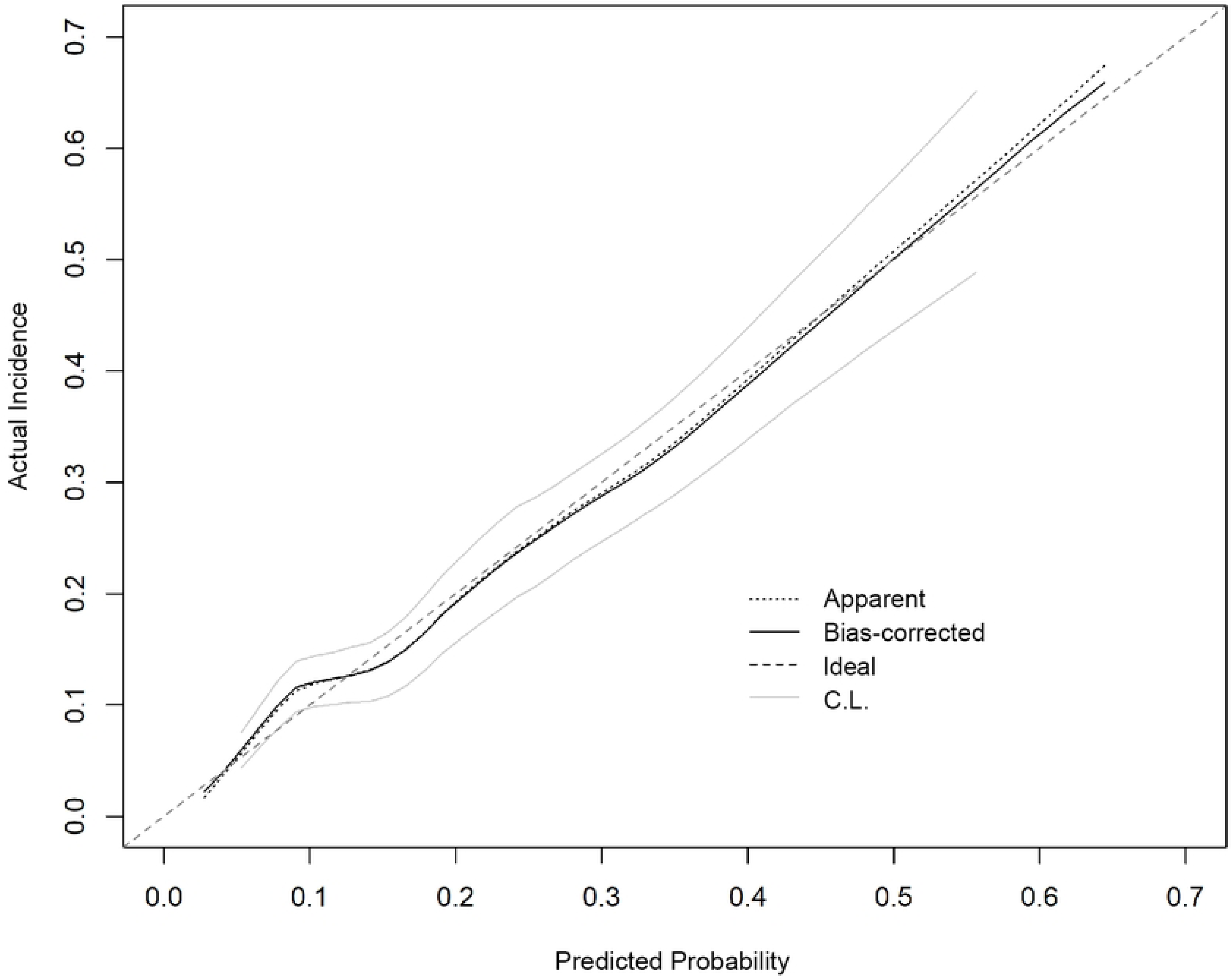
Calibration plot of the prediction model in the validation group. Apparent: The apparent calibration curve, representing the raw predictions of the model. Bias-corrected: The bias-corrected calibration curve, indicating the model’s performance after calibration. Ideal: The ideal calibration line, where the predicted probability perfectly matches the actual incidence. C.L.: Confidence Limits (or Confidence Interval), showing the range of uncertainty around the bias-corrected estimate.

### Model interpretation and clinical translation

To further elucidate the contribution of predictors and assess clinical applicability, we integrated model interpretation, decision curve analysis (DCA), and online tool development.

### Model interpretability

SHapley Additive exPlanations (SHAP) analysis was performed based on the XG Boost model to validate predictor importance from a machine learning perspective. Both the SHAP summary plot (Fig 6) and bees warm plot (Fig 7) identified duration of mechanical ventilation, antibiotic usage patterns, and urinary catheter days as the top contributors to model predictions. Importantly, the directionality and ranking of these variables were consistent with the Wald χ² statistics derived from the logistic regression model, confirming the robustness of the selected predictors across classical statistics and machine learning frameworks.

**Fig 6.**
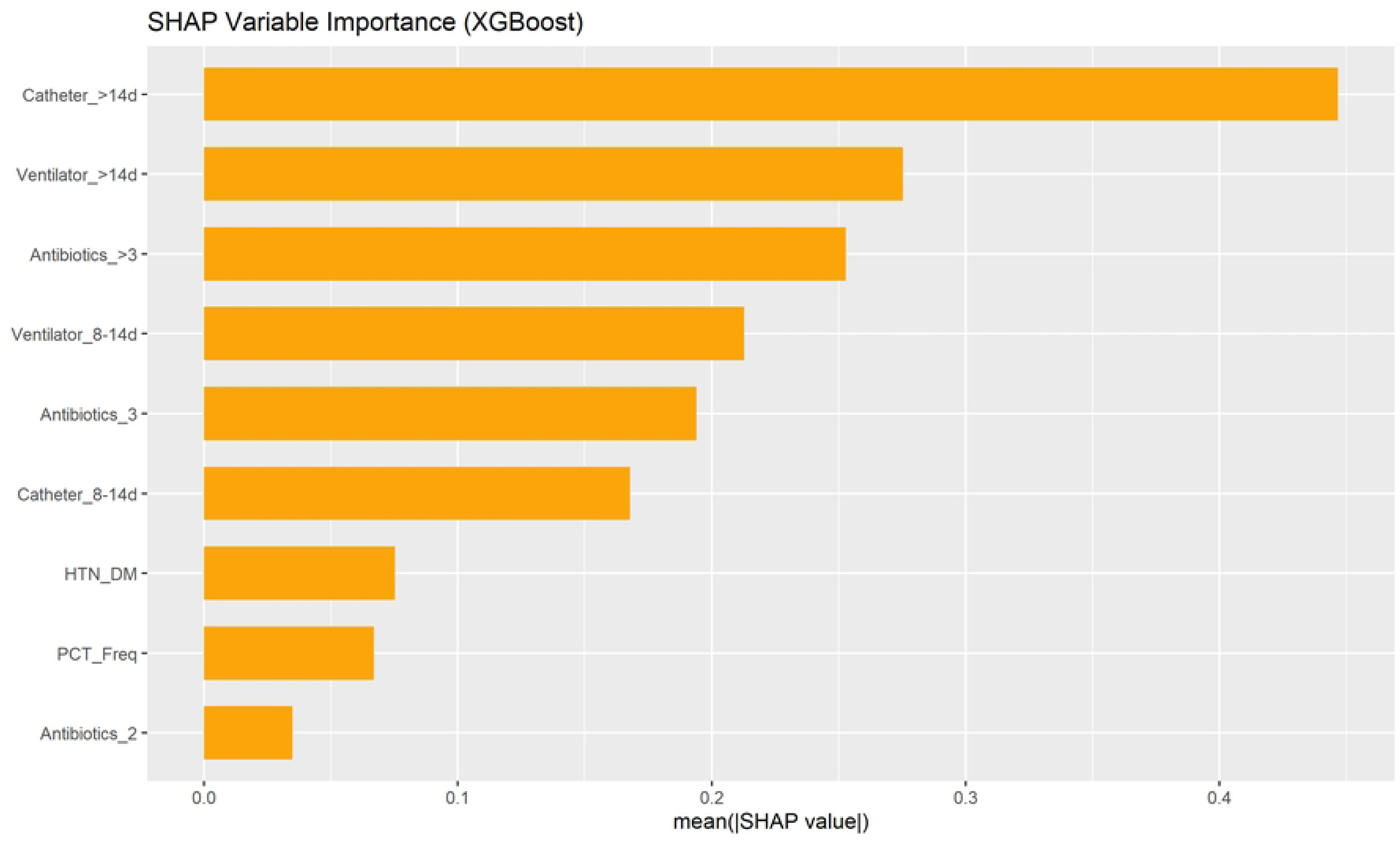
SHAP variable importance plot. Mean absolute SHAP values indicate the relative contribution of each predictor to the model output.

**Fig 7.**
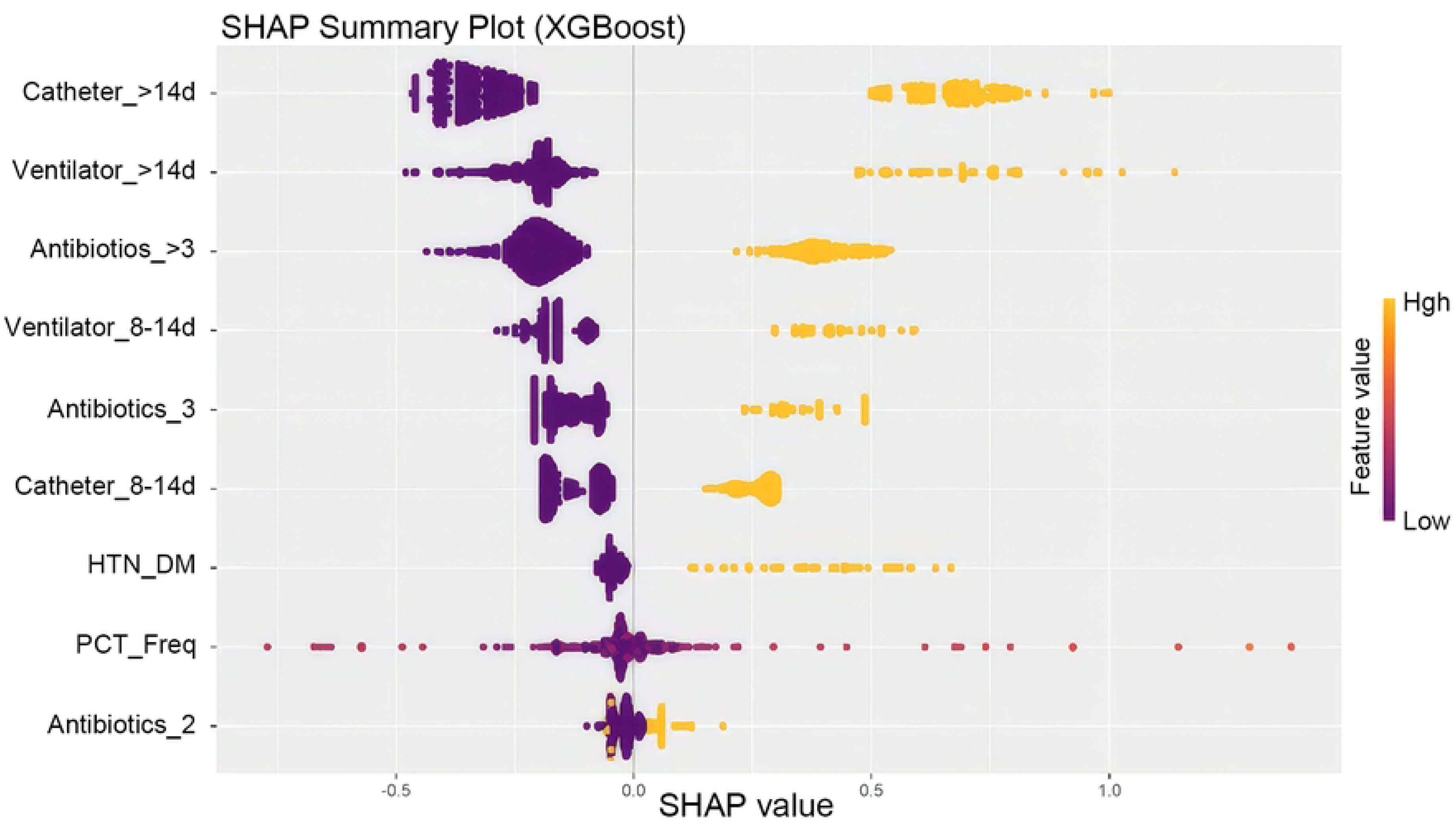
SHAP bees warm plot. Distribution of SHAP values showing the directional effect of predictors on MDR infection risk.

### Clinical utility

Decision Curve Analysis (DCA) was conducted to quantify the clinical benefit of the model (Fig 8). Across a wide range of threshold probabilities (0%–40%), the model yielded a higher net benefit than the “treat all” and “treat none” strategies, suggesting that its application in clinical decision-making would provide meaningful risk stratification without unnecessary intervention.

**Fig 8.**
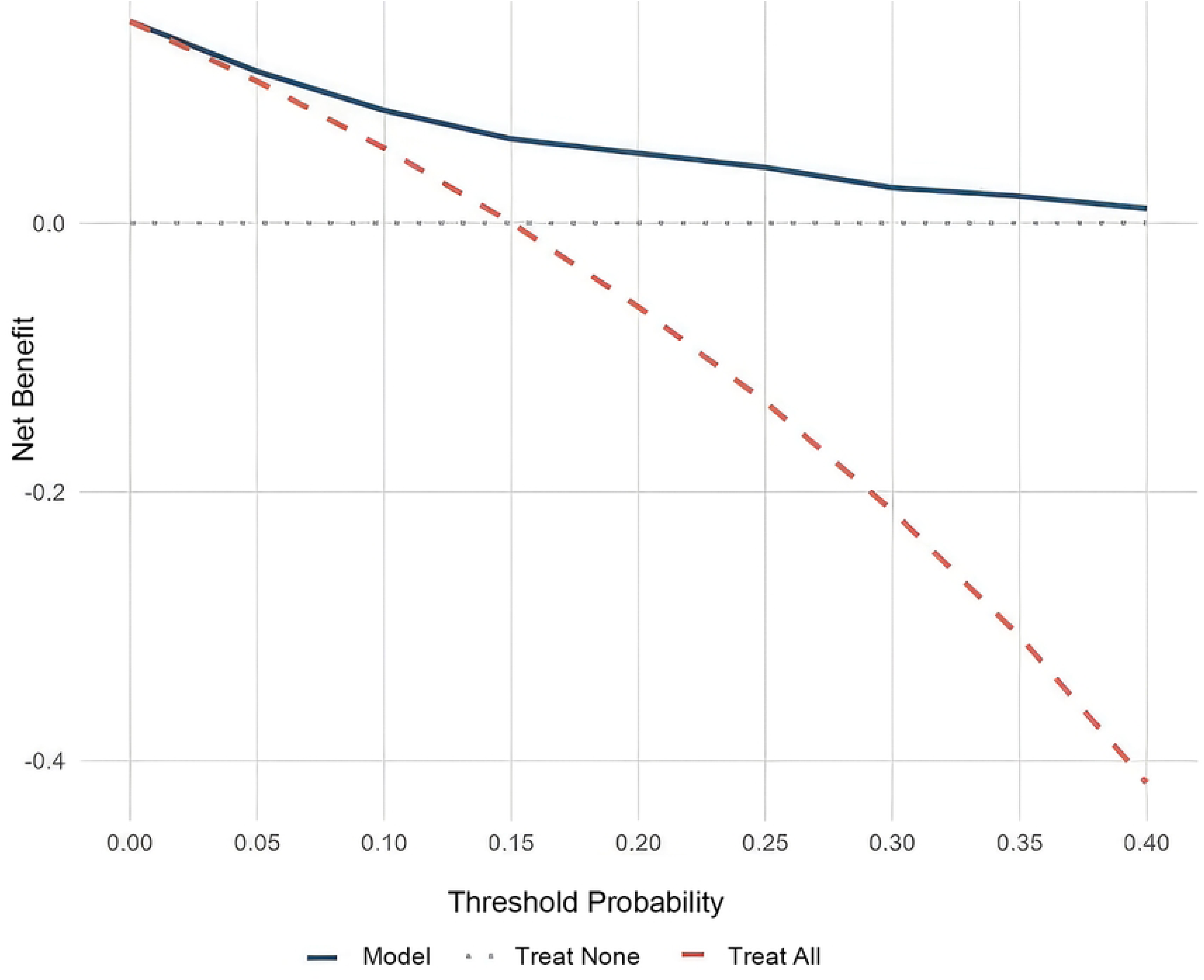
Decision curve analysis (DCA) of the predictive model. The blue solid line represents the net benefit of the proposed model. The red dashed line represents the “treat all” strategy, and the gray dashed line represents the “treat none” strategy. The model demonstrates superior clinical utility by providing a higher net benefit than both extreme strategies within the 0%–40% threshold probability range.

### Online calculator deployment

To facilitate bedside implementation, an interactive web-based calculator was developed using the R Shiny framework and deployed online (https://dongfangshao666.shinyapps.io/MDR_shiny2/). As shown in Fig 9, the interface consists of an input panel (left) for clinicians to enter five key predictors, hypertension combined with diabetes, antibiotic types, ventilator days, urinary catheter days, and PCT abnormality frequency, and an output panel (right) that dynamically generates individualized risk probabilities, nomogram visualizations, and clinical recommendations. This tool enables real-time risk stratification and supports antimicrobial stewardship at the point of care.

**Fig 9.**
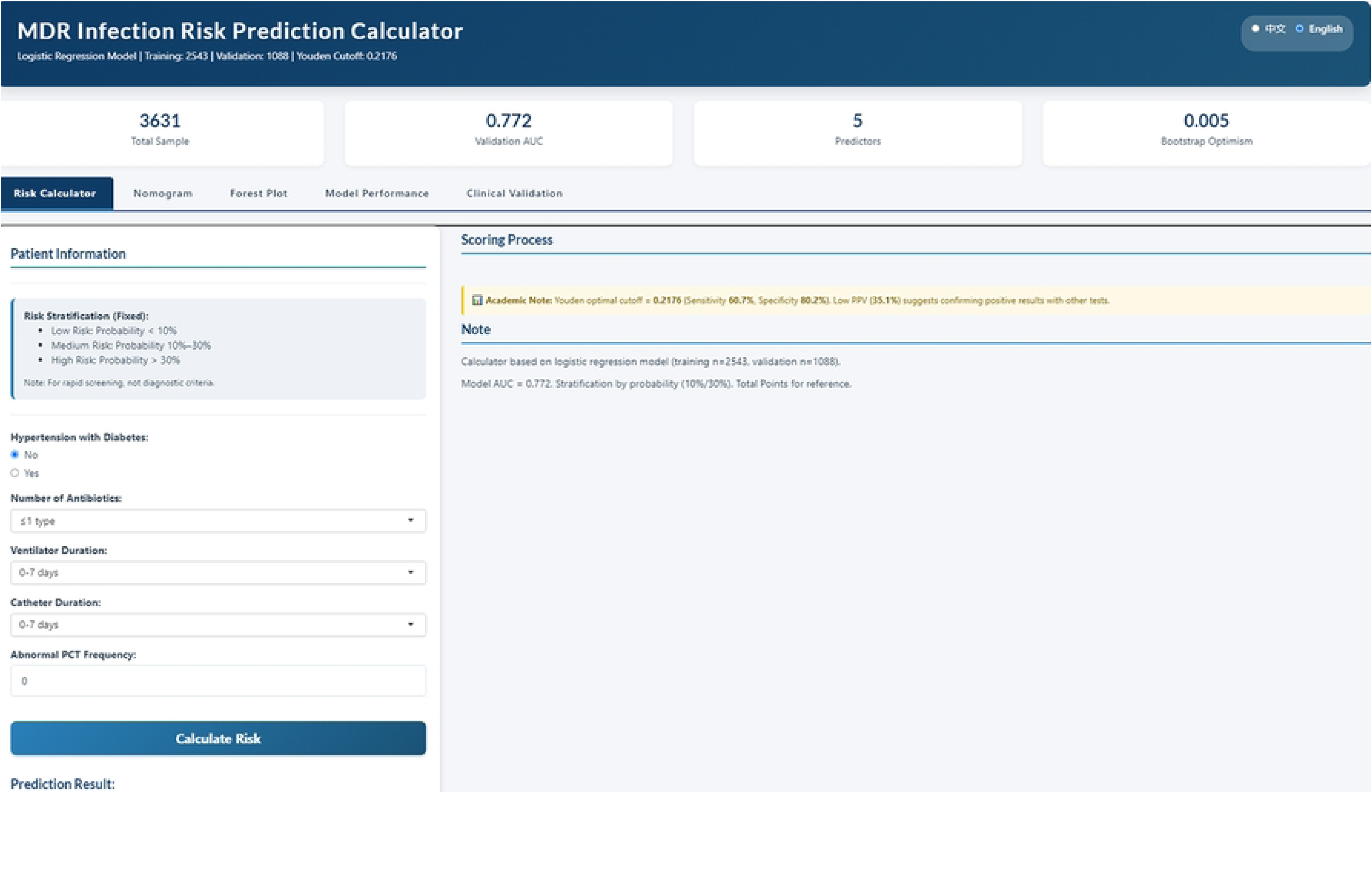
User interface of the R Shiny-based web application for real-time risk stratification for multidrug-resistant infections among ICU patients. Screenshot of the interactive MDR risk calculator deployed via R Shiny. Input interface (left panel) for five key predictors: hypertension combined with diabetes, antibiotic types, ventilator days, urinary catheter days, and PCT abnormality times. Output display (right panel) showing the individual risk probability, nomogram visualization, and corresponding clinical recommendations for bedside decision support.

## Discussion

Infections caused by multidrug-resistant bacteria in ICUs have become one of the most serious challenges facing global public health [12,13]. Due to the lack of effective antimicrobial agents, clinical treatment often faces significant challenges. This crisis of the “post-antibiotic era” is particularly acute in ICU settings, making early warning of MDR infections not merely a clinical issue, but a central concern for hospital infection control and antimicrobial stewardship [14,15].

In this study, we developed a dynamic risk prediction model using the Boruta algorithm and multivariate logistic regression. We identified five independent predictors: hypertension complicated by type 2 diabetes, concurrent antibiotic count, ventilator days, urinary catheterization duration, and the frequency of abnormal procalcitonin (PCT) readings. These factors reflect the dual nature of MDR risk, originating from both the host’s underlying pathophysiology and iatrogenic exposures within the ICU environment.

Our findings align with the current consensus that prolonged invasive device use is a primary driver of MDR acquisition [16]. Much evidence suggests that the occurrence of MDR infections results from the interaction between the host’s underlying pathological condition and specific medical exposures within the ICU. In particular, the duration of invasive procedures is widely recognized as one of the most important modifiable risk factors for MDR infections [17]. In this study, both the duration of ventilator use, and the duration of urinary catheterization were included in the final model and were associated with high odds ratios, further confirming that reducing the duration of use of these devices is a critical component of infection control. Additionally, this study revealed that combination therapy involving three or more antibiotics serves as an independent risk factor for resistance. This underscores that overly aggressive broad-spectrum empiricism, a key target for antimicrobial stewardship, can exert selective pressure favoring resistant organisms. Notably, the inclusion of the number of PCT abnormalities, rather than a single static measurement, highlights the value of dynamic monitoring in capturing the evolving risk profile of critically ill patients.

Furthermore, our results challenge the prevailing trend of pursuing algorithmic complexity. While machine learning models like XG Boost (AUC = 0.763) and Random Forest (0.703) were evaluated, the classical logistic regression model (AUC = 0.772) demonstrated superior or comparable performance. This result offers important methodological insights and suggests that in clinical prediction, “more complex” does not equate to “better”. Model parsimony and transparency are often more valuable for end-users. [18–20]. In addition, this study incorporated SHAP analysis, which not only validated the ranking of variable importance but also addressed the interpretability limitations of traditional statistical models, thereby enhancing the credibility of the results [21,22]. This not only validated the variable importance ranking but also provided a granular interpretation of feature contributions, bridging the gap between statistical inference and clinical intuition [23].

The ultimate utility of a predictive model lies in its integration into clinical workflow. Many previous models have remained at the stage of statistical charts, making it difficult to integrate them into clinical workflows [24–26]. We deployed our model as a web-based calculator via the R Shiny framework. This tool translates five routine indicators into real-time, personalized risk stratifications (low, moderate, high). This tool can effectively support clinical decision-making. For low-risk patients, it helps avoid unnecessary escalation to broad-spectrum antibiotics or excessive isolation. For high-risk patients (risk probability > 30%), it prompts early implementation of contact isolation, increased microbiological testing, or adjustments to empirical antibiotic therapy. This precision intervention strategy based on risk stratification helps optimize the use of healthcare resources while achieving infection control.

## Limitations

Several limitations should be acknowledged. First, the single-center retrospective design, although mitigated by rigorous internal validation, necessitates external validation across diverse healthcare settings. Second, while our MDR definition relied on standardized in vitro susceptibility testing (non–susceptibility to ≥3 antimicrobial classes), it did not differentiate between specific bacterial species or resistance phenotypes, potentially limiting pathogen–specific risk stratification. Finally, unmeasured confounders, including healthcare worker hand hygiene adherence and environmental contamination levels, were not included, which may have attenuated the model’s ability to capture the full spectrum of transmission–related risks.

## Conclusion

This study successfully developed a risk prediction model for multidrug–resistant bacterial infections in adult ICU patients. Based on readily available routine indicators, the model demonstrated robust predictive performance and clinical interpretability. The accompanying web–based calculator provides clinicians with a practical decision–support tool, facilitating the early identification of high–risk patients and enabling timely antimicrobial stewardship and infection control interventions.

## Author Contributions

**Conceptualization:** Lingling Ye, Wongsa Laohasiriwong.

**Data curation:** Lingling Ye, Qijuan Yang, Xiuhuan Mou.

**Formal analysi**s: Binghang Lyu, Lingling Ye.

**Funding acquisition:** Binghang Lyu, Lingling Ye.

**Investigation:** Lingling Ye, Qijuan Yang, Xiuhuan Mou.

**Methodology:** Lingling Ye, Binghang Lyu, Rajitra Nawawonganun.

**Project administration:** Wongsa Laohasiriwong, Lingling Ye.

**Resources:** Lingling Ye, Qijuan Yang.

**Software:** Binghang Lyu, Xiuhuan Mou.

**Supervision:** Wongsa Laohasiriwong.

**Validation:** Rajitra Nawawonganun, Binghang Lyu.

**Visualization:** Lingling Ye, Binghang Lyu.

**Writing original draft:** Lingling Ye.

**Writing review & editing:** Wongsa Laohasiriwong, Binghang Lyu, Rajitra Nawawonganun, Qijuan Yang, Xiuhuan Mou.

## Data Availability

The data that support the findings of this study are available on request from the first author, [Lingling Ye], upon reasonable request. According to local ethical guidelines, the use of de-identified retrospective data does not require additional IRB approval or informed consent.

## References

1. Kalın G, Alp E, Chouaikhi A, Roger C. Antimicrobial multidrug resistance: Clinical implications for infection management in critically ill patients. Microorganisms. 2023;11(10):2575. 10.3390/microorganisms11102575

2. Kreitmann L, Vasseur M, Jermoumi S, Nseir S. Relationship between immunosuppression and intensive care unit-acquired colonization and infection related to multidrug-resistant bacteria: a prospective multicenter cohort study. Intensive Care Med. 2023;49(2):154–165. 10.1007/s00134-022-06954-0

3. Su L-H, Chen I-L, Tang Y-F, Liu J-W. Increased financial burdens and lengths of stay in patients with healthcare-associated infections due to multidrug-resistant bacteria in intensive care units: a propensity-matched case-control study. PLoS ONE. 2020;15(5):e0233265. 10.1371/journal.pone.0233265

4. Martins APS, Da Mata CPSM, Dos Santos UR, De Araújo CA, Leite EMM, De Carvalho LD, Vidigal PG, Vieira CD, Dos Santos-Key SG. Association between multidrug-resistant bacteria and outcomes in intensive care unit patients: A non-interventional study. Front Public Health. 2024;11:1297350. 10.3389/fpubh.2023.1297350

5. Neuman I, Shvartser L, Teppler S, Friedman Y, Levine JJ, Kagan I, Bishara J, Kushinir S, Singer P. A machine-learning model for prediction of acinetobacter baumannii hospital acquired infection. PLoS ONE. 2024;19(12):e0311576. 10.1371/journal.pone.0311576

6. Martínez-Agüero S, Soguero-Ruiz C, Alonso-Moral JM, Mora-Jiménez I, Álvarez-Rodríguez J, Marques AG. Interpretable clinical time-series modeling with intelligent feature selection for early prediction of antimicrobial multidrug resistance. Future Gener Comp Syst. 2022;133:68–83. 10.1016/j.future.2022.02.021

7. Cao H, Oghenemaro EF, Latypova A, Abosaoda MK, Zaman GS, Devi A. Advancing clinical biochemistry: Addressing gaps and driving future innovations. Front Med. 2025;12:1521126. 10.3389/fmed.2025.1521126

8. Zhao Q, Zhang C, Zhang W, Zhang S, Liu Q, Guo Y. Applications and challenges of biomarker-based predictive models in proactive health management. Front Public Health. 2025;13:1633487. 10.3389/fpubh.2025.1633487

9. Efthimiou O, Seo M, Chalkou K, Debray T, Egger M, Salanti G. Developing clinical prediction models: A step-by-step guide. BMJ. 2024;386:e078276. 10.1136/bmj-2023-078276

10. Sharma V, Ali I, Van Der Veer S, Martin G, Ainsworth J, Augustine T. Adoption of clinical risk prediction tools is limited by a lack of integration with electronic health records. BMJ Health Care Inform. 2021;28(1):e100253. 10.1136/bmjhci-2020-100253

11. Ye Q, Chen X, Zhang J, Lin J. Meta-analysis of risk factors for infection by multi-drug-resistant organisms in intensive care unit patients. J Hosp Infect. 2025;158:1–10. 10.1016/j.jhin.2025.01.015

12. Maina JW, Onyambu FG, Kibet PS, Musyoki AM. Multidrug-resistant gram-negative bacterial infections and associated factors in a kenyan intensive care unit: A cross-sectional study. Ann Clin Microbiol Antimicrob. 2023;22(1):85. 10.1186/s12941-023-00636-5

13. Cornistein W, Balasini C, Nuccetelli Y, Rodriguez VM, Cudmani N, Roca MV, Sadino G, Brizuela M, Fernández A, González S, Águila D, Macchi A, Staneloni MI, Estenssoro E. Prevalence and mortality associated with multidrug-resistant infections in adult intensive care units in argentina (PREV-AR). Antimicrob Agents Chemother. 2023;69(3):e01426–24. 10.1128/aac.01426-24

14. Nanao T, Nishizawa H, Fujimoto J. Empiric antimicrobial therapy in the intensive care unit based on the risk of multidrug-resistant bacterial infection: A single-centre case‒control study of blood culture results in japan. Antimicrob Resist Infect Control. 2023;12(1):99. 10.1186/s13756-023-01303-2

15. Toori KU, Arshad RG, Rahim J. Comparing multidrug-resistant (MDR) and non-MDR patients in intensive care unit: Outcomes and challenges. Pak J Med Sci. 2025;41(9):2505–2511. 10.12669/pjms.41.9.12029

16. Ye Q, Chen X, Zhang J, Lin J. Meta-analysis of risk factors for infection by multi-drug-resistant organisms in intensive care unit patients. J Hosp Infect. 2025;158:1–10. 10.1016/j.jhin.2025.01.015

17. Diao H, Lu G, Zhang Y, Wang Z, Liu X, Ma Q, Yu H, Li Y. Risk factors for multidrug-resistant and extensively drug-resistant acinetobacter baumannii infection of patients admitted in intensive care unit: A systematic review and meta-analysis. J Hosp Infect. 2024;149:77–87. 10.1016/j.jhin.2024.04.013

18. Hua Y, Stead TS, George A, Ganti L. Clinical risk prediction with logistic regression: Best practices, validation techniques, and applications in medical research. Acad Med Surg. 2025. 10.62186/001c.131964

19. Christodoulou E, Ma J, Collins GS, Steyerberg EW, Verbakel JY, Van Calster B. A systematic review shows no performance benefit of machine learning over logistic regression for clinical prediction models. J Clin Epidemiol. 2019;110:12–22. 10.1016/j.jclinepi.2019.02.004

20. Pate A, Riley RD, Collins GS, Van Smeden M, Van Calster B, Ensor J, Martin GP. Minimum sample size for developing a multivariable prediction model using multinomial logistic regression. Stat Methods Med Res. 2023;32(3):555–571. 10.1177/09622802231151220

21. Stenwig E, Salvi G, Rossi PS, Skjærvold NK. Comparative analysis of explainable machine learning prediction models for hospital mortality. BMC Med Res Methodol. 2022;22(1):53. 10.1186/s12874-022-01540-w

22. Assis A, Dantas J, Andrade E. The performance-interpretability trade-off: a comparative study of machine learning models. J Reliable Intell Environ. 2025;11(1):1. 10.1007/s40860-024-00240-0

23. Luo H, Xiang C, Zeng L, Li S, Mei X, Xiong L, Liu Y, Wen C, Cui Y, Du L, Zhou Y, Wang K, Li L, Liu Z, Wu Q, Pu J, Yue R. SHAP based predictive modeling for 1 year all-cause readmission risk in elderly heart failure patients: Feature selection and model interpretation. Sci Rep. 2024;14:17728. 10.1038/s41598-024-67844-7

24. Chan SL, Lee JW, Ong MEH, Siddiqui FJ, Graves N, Ho AFW, Liu N. Implementation of prediction models in the emergency department from an implementation science perspective—determinants, outcomes, and real-world impact: A scoping review. Ann Emerg Med. 2023;82(1):22–36. 10.1016/j.annemergmed.2023.02.001

25. Lee TC, Shah NU, Haack A, Baxter SL. Clinical implementation of predictive models embedded within electronic health record systems: A systematic review. Informatics. 2020;7(3):25. 10.3390/informatics7030025

26. Jiang LY, Liu XC, Nejatian NP, Nasir-Moin M, Wang D, Abidin A, Eaton K, Riina HA, Laufer I, Punjabi P, Miceli M, Kim NC, Orillac C, Schnurman Z, Livia C, Weiss H, Kurland D, Neifert S, Dastagirzada Y, Kondziolka D, Cheung ATM, Yang G, Cao M, Flores M, Costa AB, Aphinyanaphongs Y, Cho K, Oermann EK. Health system-scale language models are all-purpose prediction engines. Nature. 2023;619(7969):357–362. 10.1038/s41586-023-06160-y

